# Clinical Usability of Generative Artificial Intelligence for MR Safety Advice

**DOI:** 10.1101/2025.10.16.25336153

**Authors:** Heather E. L. Rose, James C. Thorpe, Rafal Panek, Elsa Goncalves, Paul S. Morgan

## Abstract

This study investigated whether readily available, generative AI models, could be used to answer MR safety queries as an MR Safety Expert (MRSE), with “clinical usability” assessed by an expert review panel. This study is a mixed retrospective-prospective, proof-of-concept study. A clinical MR safety advice archive (January 2024 to April 2025) was used to curate 30 generic MR safety support requests with associated MRSE responses. ChatGPT-4o (ChatGPT) and Google AI Overview (GAIO) were prompted with these generic requests to generate AI safety advice. An expert panel assessed all answers for clinical usability. Unusable responses were assigned as; “Unsafe Advice”, “Safe but Incorrect”, “Incomplete Advice/ Key Details Missing”, “Contradictory Statements”,” Out of Date”. Requests were subcategorised into “specific” and “generic” requests, as well as “passive” and “active” implants, and “other” requests for post review analysis. Percentages of usable answers and reasons for non-usable responses were compared. Overall, 93% (28/30) of the human responses, 50% (15/30) of the GAIO responses and 43% (13/30) of the ChatGPT responses were deemed acceptable for clinical use. Subcategorization usability was: “generic”; Human 94% (16/17), GAIO and ChatGPT 59% (10/17), “specific”: Human 92% (12/13), GAIO 38% (5/13), ChatGPT 23% (3/13), Active: Human 100% (9/9), GAIO 33% (3/9), ChatGPT 22% (2/9), “passive”: Human 88% (14/16), GAIO 56% (9/16), ChatGPT 50% (8/16) and “other”; Human 100% (5/5), GAIO and ChatGPT 60% (3/5). While both AIs were able provide clinically acceptable answers for some requests they did so at a significantly lower success rate than a human MRSE.

## Introduction

Magnetic resonance imaging (MRI) plays an important role in the clinical management of patients with over 40 million(1) and 4.4 million(2) MRI scans performed each year in the US and UK respectively. Most of these scans occur without incident(3) however, MRI still has associated safety concerns with national guidelines in place to minimise risks(4,5). One area of particular concern is an increasing number of patients with implanted medical devices, often classified as either active - with a power supply for functionality, or passive - requiring no power supply(4,6). The presence of such implants or metallic foreign objects can increase the risk associated with scanning a patient (7). Additionally, the introduction of metallic objects and clinical equipment into the scanning environment can increase the risk to both patients and staff. Maximising patients access to scanning; while minimising patient and staff risk, through proportionate safety screening and scientific support, is a priority(7).

It is recommended that MRI scanning services are supported by an MR safety expert (MRSE)(4,8), with accreditation schemes(9,10) established to certify a minimum standard of expertise required. MRSEs should possess a high level of scientific and physics expertise relating to the safe operation of an MRI facility and be capable of providing high level advice on administrative, scientific, and case specific safety issues(11). Consultation with an MRSE is therefore an essential step in the patient scanning pathway and for activities within the MR environment, particularly in complex cases(4,8,11). Exploring methods that would expedite safety assessment by MRSEs could benefit both patients and workflow efficiencies.

Generative Artificial Intelligence (AI) produces new outputs from patterns learned in existing data(12) and is emerging as a potentially powerful tool for healthcare(13). Radiology is an area of prominent interest for AI application due to its complex workflows and time-consuming interpretation requirements(14,15). Specialized AI models have been successfully used to provide clinical standard answers to medical questions(16), generate reports for chest x-rays from a variety of clinical settings(17) and most relevant to the work of an MRSE, identifying patients with contradictions for MRI(18). Less specialised models, such as ChatGPT-3o, also have a place in radiology workflow optimisation, having been used to summarise radiologist reports and produce accurate and understandable patient focused equivalents(19). These generalist models, such as ChatGPT, are becoming more prominent and easily accessed by a larger pool of users(20). Additionally, Google’s AI overview (GAIO) – the AI summary generated during a google search(21) – now responds automatically on a large proportion of google searches, embedding it in people’s problem-solving processes(20,22). This study investigated whether these generative AI models could be utilised for MR Safety Expert safety queries, assessed by expert review panel.

## Materials and Methods

### Study design

This study is a mixed retrospective-prospective, proof-of-concept study, designed to explore the usability of AI to answer MRI safety queries. The use of the deidentified, retrospective datasets in this work was reviewed by Nottingham University Hospitals NHS Trust’s Caldicott Guardian, following UK’s Health Research Authority (HRA) guidelines, and determined not to require HRA ethical approval. Human MRSE responses, collected retrospectively, and AI generated responses, collected prospectively were curated and reviewed by an expert panel for “clinical usability”.

### Data Generation

#### Data sources

An existing MR safety advice archive from a large acute teaching hospital, dating from November 2018 to April 2025, was utilised for this study. MR focused medical physics support requests were logged and archived along with the responding MRSE advice. A subset of requests, dated between January 2024 and April 2025, were extracted and categorised into 18 clinically representative groupings, centred around device type or equipment support, listed in supplementary table 1.

A subset of requests from a single MRSE responder was isolated, with the corresponding MRSE responses, and ringfenced as the data source for this study. A formal power calculation was not undertaken due to the semi-qualitative nature of the review process. However, a sample size of 30 was deemed sufficient to represent the spread of requests across categories and for semi-qualitative analysis(23).

#### Data preprocessing

Requests were anonymised, condensed and then sanitised to generate generic MRSE support requests, representative of the initial requests but unrelated to a specific individual. AI prompt engineering was purposely avoided to keep requests reflective of a clinician enquiry. The full process is detailed in supplementary material 2 (S2).

#### Model selection

All AI was accessed using Microsoft edge for Business, version 137.0.3296.83 (official build, 64 bit). ChatGPT was accessed using a new account holding no previous request history. The standard/free version of ChatGPT, gpt-4o, was used with no “memory” activated, meaning previously inputted chat data did not augment the results. Specific GAIO versions were not available however all AI responses for GAIO and ChatGPT were generated on 18^th^ June 2025 to allow retrospective model identification, should the opportunity arise. Explainability is not directly assessed however incomplete responses, which could include lack of explainability in answers, were noted in the review process.

#### Response generation

The human MRSE responses were collated and saved for data cleaning. The generic MR support requests were inputted into the dedicated ChatGPT account and the google.com search bar to generate responses. In instances where an AI response was not generated, minor rephrasing would be attempted, while retaining relevant information. i.e. “a device implanted in 2005” could be rephrased to “a device implanted 20 years ago”. Further minor prompt engineering was allowed if required but noted for post review analysis. The ChatGPT and GAIO responses, without reference links, were saved offline for data cleaning. All responses, human and AI, were subjected to the same cleaning and review process as the initial requests with the additional steps listed in S2 “response cleaning”.

Data collation and cleaning was completed by an imaging scientist with 17 years’ experience then validated by an MRSE, excluded from the review panel, with 19 years’ experience. Human and AI responses were randomised for review by an expert panel.

### Panel review

We utilised an expert panel review in this study, as has been used previously when assessing generative AI in Radiology(17). Prior methodology employed direct comparison between AI and radiologist reports, with the panel members independently determining “which is better”. Our panel carried out assessment on a consensus basis with experts from a mix of radiology roles, diversifying the knowledge base. Additionally, while there is a multitude of guidance on how an MRSE should approach support requests(8,11) there is no prescribed process or template. Acknowledging that MRSE responses can vary in complexity, detail and structure while still providing a clinically usable answer, the standard of acceptance was “clinically usable”, determined by the panel.

#### Panel details and instructions

The review panel consisted of two MR clinical scientists (28 and 11 years of experience in clinical MR physics) and one senior MRI radiographer (11 years of clinical experience), who were blinded to the data preparation process and clinical context or additional case data. They were provided with the cleaned and curated dataset, plus three example safety requests for practice evaluation. Internet access, but not AI, was permitted to assist in their review process. The panel were instructed to review but not compare the responses and answer the question “Is the answer acceptable for clinical use? “Yes” or “No”. Any response deemed not acceptable for clinical use was assigned all applicable options from; “Unsafe Advice”, “Safe but Incorrect”, “Incomplete Advice/ Key Details Missing”, “Contradictory Statements”,” Out of Date”, the definitions of which are given in Table 1. Where a unanimous decision could not be reached a majority vote was used. Complete panel instructions are provided in supplementary material 3.

**Table 1:** Definitions of the justifications the expert panel could apply to responses deemed not acceptable for clinical use with examples.

| Justification | Definition | Example |
| --- | --- | --- |
| **Unsafe Advice** | Advice that could result in the patient being harmed due to the MRI scanner. | Recommending scanning an MR Unsafe implanted device / Providing unsafely incorrect scanning conditions. |
| **Safe but incorrect advice** | Advice that would not cause harm to the patient but features incorrect information. | Providing incorrect but safe scanning conditions (e.g. only 1.5T for a device that can be scanned at 1.5T or 3T) / Giving correct safety advice but incorrectly asserting that there will be substantial artifact. |
| **Incomplete advice/ Key details missing** | Insufficient information provided in the responses for it to be of clinical value. | Omitting certain safety conditions / Advice that is too generalised to be clinically useful. |
| **Contradictory statements** | Responses that contain multiple statements that contradict each other (not statements that contradict the query). | Stating a device can always be scanned safely and can never be scanned safely. |
| **Out of Date Advice** | Responses that contain advice that based on information that has been superseded. | Stating that no pacemaker can be scanned safely. |

### Subcategorization

Due to the high granularity of the pre-processing categories, requests were subcategorized into “specific”, where the make/model is provided e.g. Boston Scientific Emblem S-ICD A219, and “generic”, e.g. “conditional pacemaker” requests, for post-review analysis. A second subcategorization, where foreign body and equipment requests, labelled as “others” and were separated from implant requests, divided into “passive” and “active” implants, following the definitions outlined in local national guidelines(4), was also utilised.

### Statistical analysis

Percentages, rounded to two significant figures, for usable answers and reasons for non-usable responses from the human and AI were compared. It is important to note that multiple reasons for an “unusable” response were allowed and are reflected in the percentages. All data collation, storage and analysis were carried out in Microsoft Excel.

Variability in request types was ensured by selecting from across all usable categories in the original dataset. Responses are limited to one human responder however this was considered acceptable for a proof-of-concept study to ensure that all panel members were blinded to requests prior to review.

## Results

### Data generation

MR medical physics support requests dated between January 2024 and April 2025, totalled 141 across 18 categories of advice types provided the study data (Figure 1). Three human MRSEs were individually or jointly responsible for the MRSE responses. Two of these MRSEs were allocated to the review panel and all requests they were solely or jointly involved in removed (48). This included queries categorized as “multifaceted” which were deemed too complex to sanitise, reducing the categories to 17.

**Figure 1.**
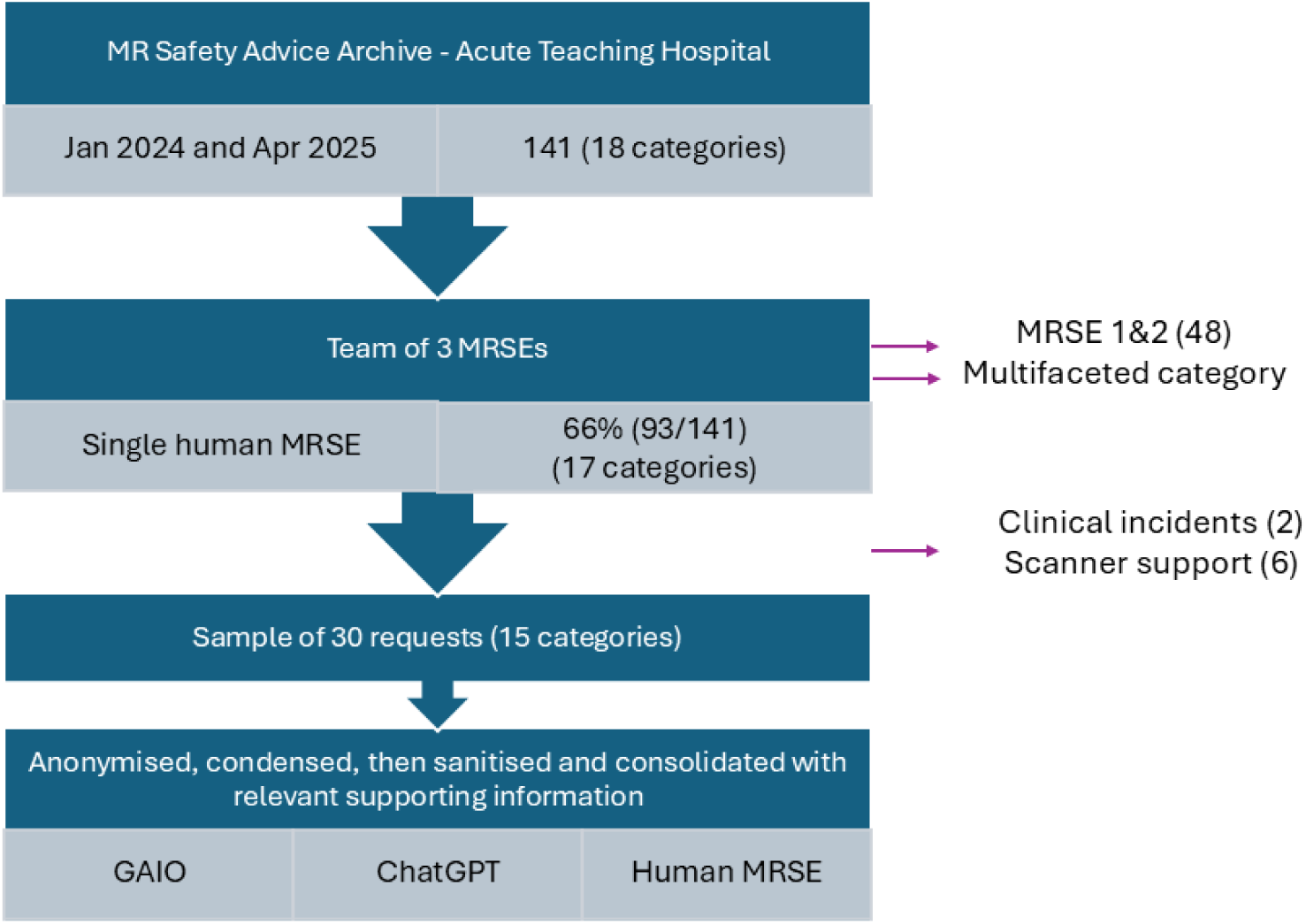
MR safety request data flow with number of requests at each stage, data exclusion and number of request categories with final data generations from a Human MR safety expert (Human), Google AI Overview (GAIO) and ChatGPT-4o (ChatGPT).

This left one responder providing 66% (93/141) of the responses independently. Two requests relating to specific clinical “incidents” and six “scanner” requests focusing heavily on local policy were removed, leaving 15 categories. A representative subset of 30 queries were selected across the 15 categories.

All required data was available to consolidate the clinical requests into representative generic MRSE requests and to condense all human responses. No data loss occurred at this stage. There were four occasions prompt engineering was required for GAIO to initiate. No prompt engineering was required for ChatGPT however for one query ChatGPT provided two responses, one of which was randomly selected for panel review.

### Panel review

The panel were unanimous in their decision for 84 out of the 90 responses provided. A majority vote was used in the remaining 6 cases.

93% (28/30) of the human responses, 50% (15/30) of the GAIO responses and 43% (13/30) of the ChatGPT responses were deemed “acceptable for clinical use” (Figure 2). AI responses, irrespective of final decision, were frequently labelled as “overly verbose”.

**Figure 2.**
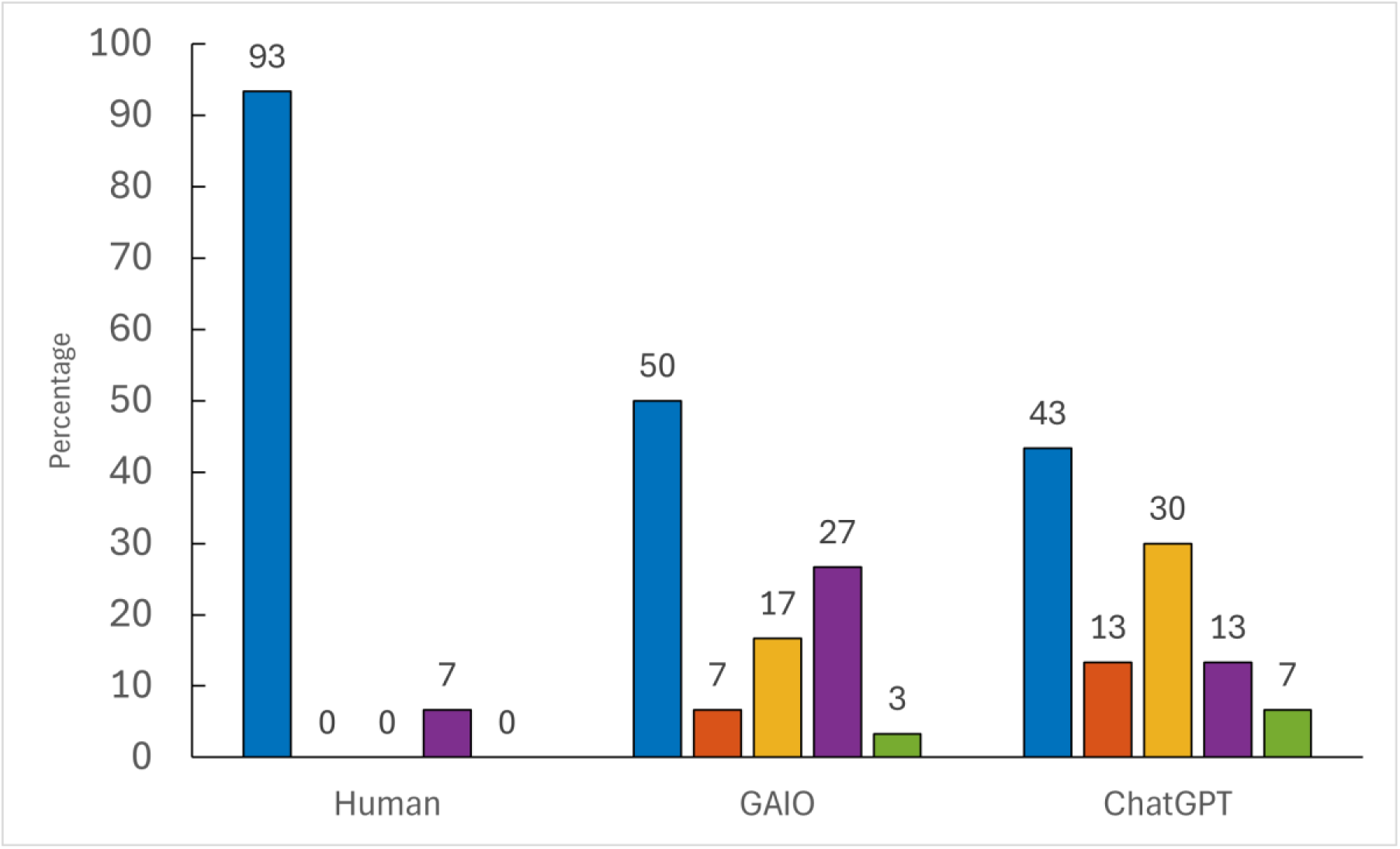
Percentage of responses that were accepted by the panel as clinically usable (blue), or rejected for the non-exclusive reasons of “Unsafe Advice” (orange), “Safe but Incorrect Advice” (yellow), Incomplete Advice/Key Details Missing” (purple), and “Contradictory Statements” (green) for the Human MR safety expert(Human), Google AI Overview (GAIO) and ChatGPT-4o (ChatGPT)

### Clinically un-usable responses

Clinically unusable results are detailed in Table 2 with full comments in supplementary table 4. One request concerning three, overlapping, stents was deemed unusable in all three responses. The other unusable human answer was later determined to be due to a typo changing the meaning of the response. 11 of the 21 unusable responses were usable from both AIs. One GAIO response was considered both [“Unsafe Advice” and “Contradictory Statements”]. For ChatGPT one response was [“Safe but Incorrect” and “Incomplete advice/Key details missing”] and one [“Unsafe Advice” and “Contradictory Statements”].

**Table 2:** Breakdown of reasons given by the expert panel for responses deemed unacceptable for clinical use. Responders are abbreviated to Human MR safety expert (Human), Google AI Overview (GAIO) and ChatGPT-4o (ChatGPT).

| **Responder** | **Unsafe Advice** | **Safe but Incorrect** | **Incomplete advice/Key** | **Contradictory statements** | **Out of Date** |
| --- | --- | --- | --- | --- | --- |
| **Human** | - | - | 7% (2/30) | - | 0% |
| **GAIO** | 7% (2/30) | 17% (5/30) | 27% (8/30) | 3% (1/30) | 0% |
| **ChatGPT** | 13% (4/30): | 30% (9/30) | 13% (4/30) | 7% (2/30) | 0% |

### Subcategorization

Requests were subcategorised firstly into generic (17) and specific (13), as defined in the materials and methods section. Human responses were usable for 94% (16/17) of generic requests and 92% (12/13) of specific requests. GAIO and ChatGPT both provided usable answers for 59% (10/17) of generic request but only 38% (5/13) and 23% (3/13) of specific requests respectively (Figure 3a and 3c). A second subcategorization of active implants (9), passive implants (16), and other/ nonimplant requests (5) found that the human responses where usable 100% of the time for both active (9/9) and other (5/5) and 88% (14/16) for passive implants. Both GAIO and ChatGPT provide usable answers for 60% (3/5) of “other” requests (Figure 3b and 3d). For passive implants GAIO and ChatGPT answers were usable 56% (9/16) and 50% (8/16) of the time respectively. AI performed worst on requests for active implants with 33% (3/9) usability for GAIO and 22% (2/9) for ChatGPT. A breakdown of the reasons for un-usability for all subcategorises can be found in Figure 3.

**Figure 3.**
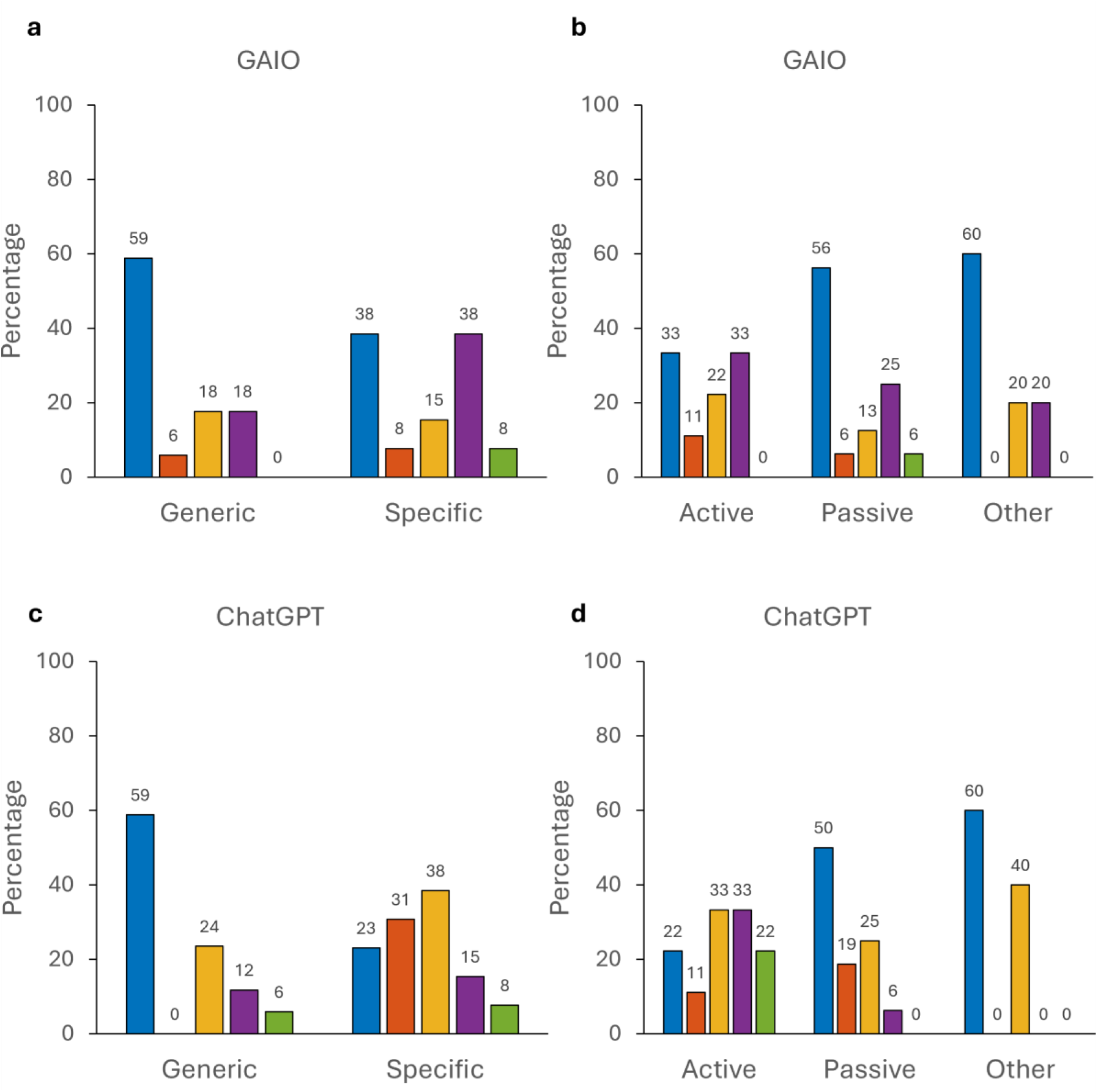
Percentage of responses by subcategory that were accepted by the panel as clinically usable (blue), or rejected for the non-exclusive reasons of “Unsafe Advice” (orange), “Safe but Incorrect Advice” (yellow), Incomplete Advice/Key Details Missing” (purple), and “Contradictory Statements” (green) for (a) Google AI Overview(GAIO) subcategorised into “generic” and “specific” requests, (b) GAIO subcategorised into “Active”, “Passive” and “Other” implant types, (c) ChatGPT subcategorised into “generic” and “specific” requests, and (d) ChatGPT subcategorised into “Active”, “Passive” and “Other” implant types.

## Discussion

An MR safety query archive was utilised to produce a dataset of clinically representative MRSE queries, with human expert responses. These queries were used to prompt GAIO and ChatGPT and these and the human responses were evaluated for clinical usability by a panel of experts. The human responses performed best with a 93% usability, followed by GAIO (50%) and then ChatGPT (43%). No answers produced by the human were found to be “Unsafe Advice”. AI performed best on passive device and “other” equipment requests and more generalised requests, while still falling behind the level of clinical usability reached by a human MRSE.

Among MR professionals there is a high level of knowledge and a willingness to accept AI into clinic practice(24) however, AI should enhance the clinical workflow while maintaining clinical quality and abiding to ethical and legal requirements(25,26). To the authors knowledge there have been no direct evaluations of AI use for MRSE support requests. Several Natural Language Processing (NLP) algorithms have been used to identify patients with contradictions to MRI from medical records(18). Both expert informed, and ontology-derived models were able to identify patients with higher risk devices with an accuracy of over 80% but did not consider whether AI can provide safety advice after identification.

This study showed the human MRSE achieved a high standard of clinical acceptability reflective of the experience, level of training and evidence-based assessments an MRSE undergoes(9,10). Both GAIO and ChatGPT performed at a much lower level of clinical acceptability. Differences have previously been found in how specialists and generalists clinicians perform on various clinical tasks(27). Clinical specialists were less risk adverse with treatment decisions and more up to date on relevant clinical developments than generalist practitioners, who were found to be more risk adverse and less likely to shed out of date practices and accept new advances. We found that ChatGPT aligned with the risk adverse nature of generalist clinicians, being most likely to be clinically unusable due to being “safe but incorrect”. While superficially this can seem the preferable option for AI failure this includes recommendations to “Not Scan” when the patient could undergo an MR scan, benefiting from its diagnostic information. The poorer performance of both AIs in active and specific MRSE requests, generally assumed to be more complex, also showed a reduced level of specialist knowledge in comparison to a human MRSE. ChatGPT systematically performed worst on “specific” requests, accounting for all “unsafe” answers, showing a difficulty in providing safe clinical advice when working on the most specialized cases. GAIO behaved similarly and while more able to correctly answer “specific” requests was most likely to provide “incomplete” answers for active and specific request demonstrating an inability to fully consider complex requests. This trend of AI performing worse with more complex, specialist problems has been seen in other clinical specialisms(16). Prompt engineering may improve AIs performance however this moves us away from requests that are reflective of current clinical practice and expertise.

A possible way to successfully leverage AIs ability to gather data while acknowledging its lower clinical usability is to combine human MRSE expertise and AIs research capabilities. Workflows have previously been developed to demonstrate the potential of clinician-AI collaboration. AI generated radiology reports were successfully edited by clinical experts, improving the clinical acceptability(17) and AI has also been utilised to summarise radiologist reports, producing patient focused reports with a high-level of accuracy and significantly improved comprehension(19). This study featured an instance where GAIO highlighted potential MRI induced auditory effects in patients with cochlear implants(28). This was a phenomenon the panel was unaware of, demonstrating the potential for AI/human collaboration. By using AI as a resource for gathering links to device conditions and guidelines and providing a first draft for MRSEs, a collaborative approach may improve efficiency and expand expert knowledge. While bespoke AI models, specifically trained, may perform better in relevant scenarios(16–18) these models are less widespread and not immediately available in the same way as ChatGPT and GAIO. These models are also trained on defined medical text where MRSE requests vary greatly in nature, requiring strong technical knowledge and good clinical practice, combined with disparate device information(8,11). This need for a vast amount of specified device knowledge, that could not currently be fully accounted for in a specialist model, supports the use of these generalizable AI, initially as a data sourcing mechanism. We propose this study also provides a methodological framework that could be replicated in other clinical fields providing similar services, offering a robust and fair approach to AI evaluation.

### Limitations

The data used in this study is single centre, single human responder with a review panel from the same centre. Future the work should include multi-centre, multi responder responses with an external review. However, since the completion of our own review GAIO has been expanded with “AI mode” providing a search summary akin to the interactive nature of ChatGPT. Additionally, Open AI, the providers of ChatGPT, have introduced ChatGPT-5o. The fast-moving developments and releases in the field of AI warrant accelerated research approaches to maintain pace with progress and keep users informed. The use of a historic MRSE archive means that human and AI responses gathered had to be approached with variations in methods. Future studies would include multiple MRSEs answering bespoke MRSE requests in sync with AI. This study focused on easily accessible generalisable AI however healthcare specific AI are available and could be evaluated in future work. It should be noted that both AI systems used in this study provide disclaimers about the accuracy and potential for incorrect answers. We did not assess the stability of AI answers however the framework could be modified to do so. Response stability, as models developed, should also be assessed in future work.

We have investigated the clinical usability of generative AI freeware answering Magnetic resonance safety requests. The human responses performed best with a 93% usability, followed by GAIO (50%) and then ChatGPT (43%). No human answers were found to be “Unsafe” but both AIs did provide some “unsafe” responses. While both AIs were able provide clinically acceptable answers for some MRSE requests they did so at a significantly lower success rate that a human.

## Data sharing statement

Data generated or analysed during the study are available from the corresponding author by request

## Declaration of Conflicting Interest

Heather E. L. Rose would like to declare share options in Healx Ltd. The other authors have no conflicts of interest to disclose.

## Data Availability

Data generated or analysed during the study are available from the corresponding author by request

## Notes

### Funding Statement

None

### Author Declarations

Caldicott Guardian & Chief Clinical Information Officer of Nottingham University Hospitals NHS Trust waived ethical approval for this work. Deidentified, retrospective datasets.

