## Supplementary materials for "Clinical Usability of Generative Artificial Intelligence for MR Safety Advice"

Supplementary Table 1: Definitions of the initial MR safety request categories with categories in red those that were not represented in the final analysis.

| **MR safety requests categories** |
|---|
| Clips and staples |
| Coils & EVARS |
| Stent |
| Vagus Nerve Stimulator |
| Cardiac Implantable Electronic Device |
| Cochlear Implant |
| Foreign object |
| Optometry implant |
| Gastric/weight loss |
| Equipment |
| Prosthesis |
| Scanner |
| Incident |
| Other nonactive implant |
| Other active implant |
| ExFix |
| Orthopaedic implant |
| multifaceted |

Supplementary Material 2

### MRSE request pre-processing method

1. Removal of any identifying details relating to a patient or staff member in the request information.
2. Collating request information i.e. initial support request plus any additional clarification and details and any information extracted from other clinical systems or reviewed images i.e. device size or shape as measured by review of CT.
3. Condensing and sanitising request details into a generic MR support request ensuring all information provided to the human MRSE is included but that information, clinical and non-clinical, irrelevant to the specific MR request is removed.
    a. All conversational tone or reference to other clinical systems was removed
    b. No information required to assess the MR risk associated with the request was removed
    c. Any clinical contextual information unrelated to MR risk or mitigation strategies was removed.
    d. Typos were not corrected or regional scientific terms relating to MR were not changed but English spelling was used when cleaning all data.
    e. References to local procedures were not included but the information was paraphrased if required to provide a response.
    f. All relevant information was included but effort was made to not engineer the request in to prompts designed for AI. Ultimately for the request to be representative of a layman regarding AI prompt engineering.

### Response cleaning

1. Cleaned of any repetition unless contradictory statements included.
2. General MR knowledge that is unrelated to the MR request removed.
3. For the AI responses, generic AI phrases and obvious AI formatting or picture icons were removed.

Supplementary Material 3

| Panel Instructions | Points to Note |
|---|---|
| 1. On each sheet you will find a sanitised MRI related safety question labelled "**request**" | ❶ Both requests and responses (human and AI) have been condensed and sanitised. |
| 2. You will then find responses 1, 2 and 3 - provided by a two AI "chat-bots" and one human, randomly allocated. These are sanitised AI and human answers to the detailed "request". | ❷ Information from several emails and knowledge from image reviews has been combined to construct the requests and ensure the AI and the human had the same information when providing an answer. This will make some requests appear oddly worded. The focus should be the information provided, and the advice requested, not the nuances of how it is phrased. |
| 3. For each response discuss and decide **as a panel** if the response answers the request to a clinically acceptable standard and "Yes" or "No" to the question "is the answer acceptable for clinical use?" | ❸ Responses have been condensed in the same way, combining multiple emails, links and data sources from a human responder or AI. Do not consider whether the answer is "professionally worded" but whether the information provided is correct. |
| 4. Tick "Yes" if it is and "No" if it isn't | ❹ Best efforts a have been made to randomise the AI and humans responses and remove phrases indicating the responses source. While it is difficult, don't try and figure out "who" is providing the answer. |
| 5. **If** you select **"No" chose** the option that reflects the reason you chose "**No**" - you can select more than one | ❺ Assess each response individually and do not compare answers to find a "best". An answer to the minimum clinical standard is still clinically usable even if the other answers are of a higher standard - you can however make a note if you think that is a dramatic difference. |

| | |
|---|---|
| 6. There is a "Notes" section - please only use notes INSTEAD of the given options for "**No**" if you can in no way can explain your decision using the options provided. While not required the Notes section can be used to give details of difficult decisions or to highlight big discrepancies between the responses. - **Do note if a decision is not unanimous** | ❻ Best efforts have been made to include all relevant information from the raw responses however the validity or correctness of the answers has not been checked. You may need to go look up MR conditions to confirm details are correct. |
| **Note: The Request ID for the collating scientist, should be used for the retrospective review of a response or request be required. You are not required to do anything with this information** | ❼ Some of the responses are very detailed and you, may not see all the information without expanding the formula bar. **Make sure you can see the full response when accessing it** |
| Practice questions | **Three practice questions have been included for you to test your panel processes and get a feel for the marking document. They are made up questions to simulate the review process. Please record your answers/ decisions as you would with the final quires. Make any note you wish.** |

Supplementary Table 4: A list of all requests with a clinically unusable responses, from any responder, and the reviewer's justification comments for the AI or human that provided an unusable answer.

| Request ID | Google Overview | ChatGPT | Human |
|---|---|---|---|
| **SICD off Label** | Suggests it is conditional but does not recommend referring to conditions. | Gives no reference to the conditions/conditional nature of the device. If the scan is deemed not urgent then this would imply there are no further limitations after 6 weeks which is missing important information | |
| **VNS inactive** | Mistakes VNS being off/battery depleted as required conditions rather than concerns. This is highlighted where it advices ensuring the battery is depleted before scanning. | Recommends using contrast enhanced MRI if you cannot use MRI | |
| **Mitra clip** | States that the clips are MRI safe and MRI conditional and does not specify the conditions or that they need checking + Contradictory | Conditions are incorrect and states that SAR<4W/kg is normal mode which is incorrect. Correctly identified name of document for source (despite then being wrong) | |
| **Overlapping Stents** | Unclear about whether there were safety considerations or not. The response is very generalised without answering the question | Wrong conditions for Elluvia stent. | Good but does not refer to the third stent. As the request specifically mentioned three stents it was deemed an omission of information to not mention this. |
| **SCS** | Confused by r.f. coil type - particularly transmit versus receive. Too much information given with no conclusion. Seemed to struggle with identifying the important aspects of the query | Confused by r.f. coil type - particularly transmit versus receive. Too much information given with no conclusion. Seemed to struggle with identifying the important aspects of the query | |

| | | | |
|---|---|---|---|
| **Foreign body** | Implies safety based on assumptions around unknown location of fragments | Makes incorrect statements about the nature of steel (iron-based steel is all steel), "distance from isocentre reduces risk" is incorrect. Some statements confusing e.g. "Patient is asymptomatic" | |
| **Epidural Lines** | States you cannot scan but the size of the spring is very small and can be restrained by a sandbag | States you cannot scan but the size of the spring is very small and can be restrained by a sandbag | |
| **Cardiac Mesh** | Not unanimous - 2 reviewers wanted more information about make/model (1) and conditions (1) with the term "standard conditions" deemed too ambiguous | | |
| **3T Halo Traction** | Does not provided information required to answer question. Too many subjectives | Says scan unsafe and should not happen when it can | |
| **Punctum Plug** | Implies silicon is slightly magnetic and states could cause artefact | | |
| **Gastric Band** | Did not mention required spatial gradient limit | Did not mention required spatial gradient limit and had an unexplainable reference to a 10cm distance + Incorrect | |
| **CI** | Does not specify the conditions although does suggest finding out for oneself. It also mentions "fever is a contraindication" which is doesn't make sense and that "patients may experience auditory sensations" which is also incorrect | Says PET/MRI could be used instead of MRI + Contradictory | |
| **CI under GA** | Implies that the freely rotating magnet would mean no artefact however this is not the case, there will definitely be artefact | | |
| **Unknown MCA stent** | Overly cautious and does not provide options to move the case forwards. This would mean no scan when there could be scan | | Not unanimous - confusion over "coronal stent" - this term does not exist. As the following advice is all dependent on their being no evidence for delay relating to these stents and the |

| | | | |
|---|---|---|---|
| | | | terminology unclear it is unclear as to whether the rest of the evidence is based on a term that doesn't exist |
| **IVC filter** | Does not refer to the Bird's nest filter which is the only slightly ferromagnetic filter and is considered safe at 1.5T but unknown at 3T. Imply a wider range of conditions out there and suggests scanning on 3T may be safe | Does not refer to the Bird's nest filter which is the only slightly ferromagnetic filter and is considered safe at 1.5T but unknown at 3T. Imply a wider range of conditions out there and suggests scanning on 3T may be safe | |
| **Stent** | | Incorrect conditions (no reference to decreased SAR of 1W/kg below umbilicus). | |
| **MSK metalwork** | | Defines titanium implant as MRI-safe however MRI-safe includes non-metallic in definition. Not unanimous | |
| **VNS 3T** | | States scanning cannot happen without a Tx/Rx coil which is not the case | |
| **Aneurysm Clip Old** | | Incorrect use of MR safe (referring to metallic object as potentially MRI safe) | |
| **Non Conditional CIED** | | Gives no reference to continuous monitoring during scanning implying that the scan would have no issues if the conditions followed. However, it is possible for issues to happen due to the pacemaker being out of threshold | |
| **Paed growing Prosthesis** | | States that the patient can have a scan, but the device is deemed unsafe | |
